# A Novel Retinal Vascular Feature and Machine Learning-based Brain White Matter Lesion Prediction Model

**DOI:** 10.1101/2021.09.27.21264168

**Authors:** Alauddin Bhuiyan, Pallab Kanti Roy, Tasin Bhuiyan, Elsdon Storey, Walter P Abhayaratna, Mandip Dhamoon, R. Theodore Smith, Kotagiri Ramamohanarao

## Abstract

White matter lesion (WML) is one of the common cerebral abnormalities, it indicates changes in the white matter of human brain and have shown significant association with stroke, dementia and deaths. Magnetic resonance imaging (MRI) of the brain is frequently used to diagnose white matter lesion (WML) volume. Regular screening can detect WML in early stage and save from severe consequences. Current option of MRI based diagnosis is impractical for regular screening because of its high expense and unavailability. Thus, earlier screening and prediction of the WML volume/load specially in the rural and remote areas becomes extremely difficult. Research has shown that changes in the retinal micro vascular system reflect changes in the cerebral micro vascular system. Using this information, we have proposed a retinal image based WML volume and severity prediction model which is very convenient and easy to operate.

Our proposed model can help the physicians to detect the patients who need immediate and further MRI based detail diagnosis of WML. Our model uses quantified measurement of retinal micro-vascular signs (such as arteriovenular nicking (AVN), Opacity (OP) and focal arteriolar narrowing (FAN)) as input and estimate the WML volume/load and classify its severity. We evaluate our proposed model on a dataset of 111 patients taken from the ENVISion study which have retinal and MRI images for each patient. Our model shows high accuracy in estimating the WML volume, mean square error (MSE) between our predicted WML load and manually annotated WML load is 0.15. The proposed model achieves an F1 score of 0.92 in classifying the patients having mild and severe WML load.

The preliminary results of our study indicate that quantified measurement of retinal micro-vascular features **(AVN, OP and FAN)** can more accurately identify the patients who have high risk of cardio-vascular diseases and dementia.

## Introduction

The retina is the inner surface of the eye which covers the largest part of the fundus. The main structures of a retina include the optic disc (i.e., optic nerve), the blood vessels (arteries and veins), and the macula. Retinal images are captured using a fundus camera, Figure 1 shows a color fundus image of the human retina. Retinal pathologies includes different types of retinopathy signs, arteriovenous nicking (AVN), focal arteriolar narrowing (FAN), generalized arteriolar narrowing, venular dilation, etc.; detail descriptions of these retinal pathologies are present in [1] and [2]. Retina is an extension of the inter-brain [3] and retinal micro-vasculature share anatomic, embryologic and physiologic characteristics with the cerebral or brain microvasculature [4]. Association between retinal microvascular changes and different types of cerebral abnormalities (such as stroke and dementia) have been reviewed in [5-7]. Table. 1 shows the association between retinal and brain imaging abnormalities or pathologies.

**Figure 1.**
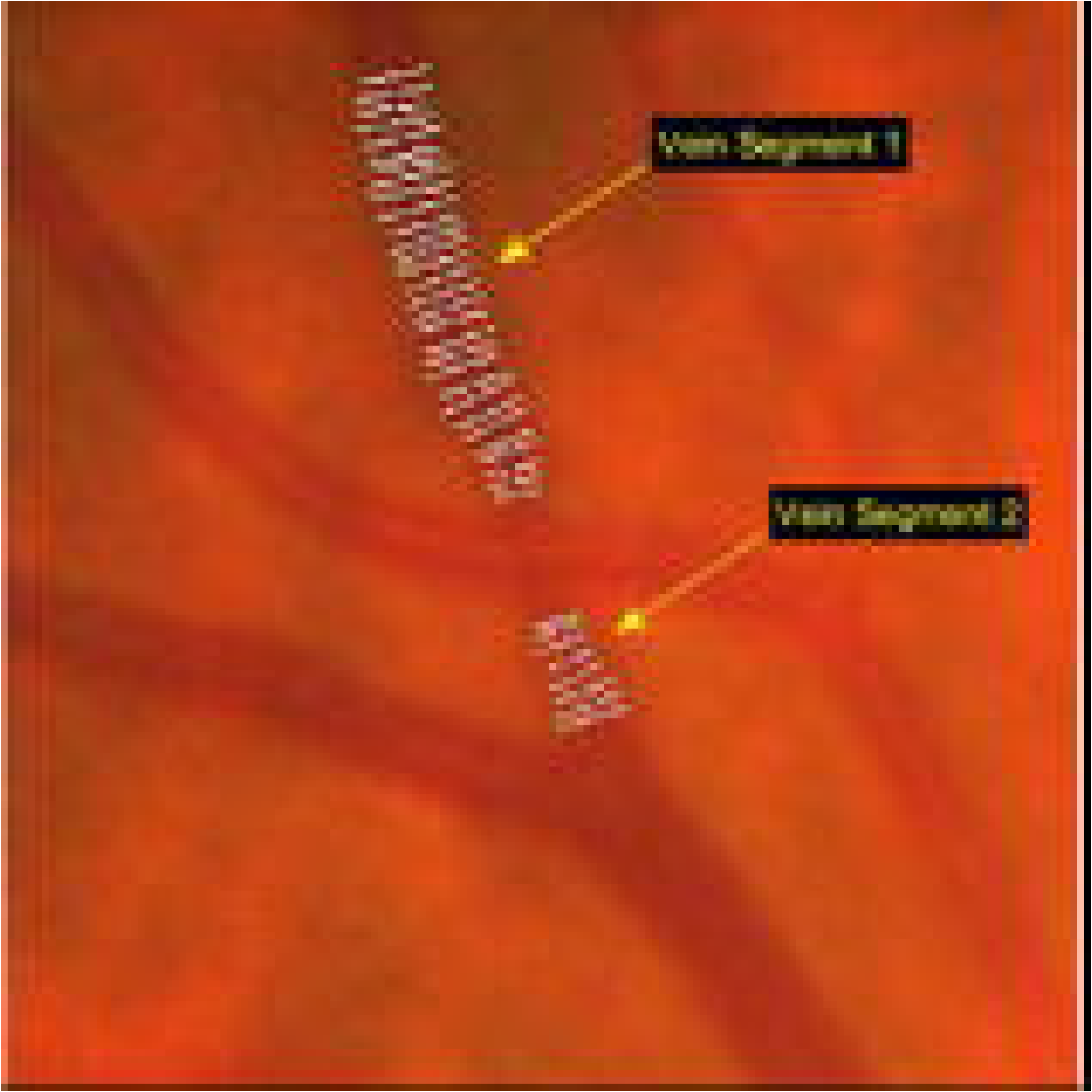
A color fundus retinal image showing different anatomical structure of retina.

Three largest population-based studies: AGES [8], ARIC [9] and CHS [10] have reported the association of retinopathy signs and retinal microvascular abnormalities with brain imaging abnormalities. Among them, only ARIC found significant association between retinopathy signs and brain imaging abnormalities which includes infarcts, WMLs and atrophy. Associations of arteriolar narrowing and venular dilation with the brain imaging abnormalities were generally weaker as reported by two population-based studies as shown in Table 1. In contrast to that, all three population-based studies [8, 10] have reported that AVN and FAN have statistically significant association with the brain abnormalities. Therefore, among different retinal pathologies, we mainly focus on the AVN and FAN for designing a prediction model of WML load. Among different brain abnormalities, we select WML because WML has a strong association with stroke, dementia and deaths as shown in Table 2.

**Table 1.** Summary of the associations between retinal imaging pathologies and brain imaging pathologies [4] based on cross-sectional population based studies.

| Retinal imaging | Study Name | Brain Imaging pathologies |
| --- | --- | --- |

| pathologies |  | Odds Ratio (95% CI) |  |  |  |  |  |
| --- | --- | --- | --- | --- | --- | --- | --- |
|  |  | Any<br>Infarct | Lacunar<br>Infarct | Subcortical<br>WML | Periventricular<br>Subcortical<br>WML | atrophy | Cortical<br>Atrophy |
| Any retinopathy | AGES [1, 2] | 1.06<br>(0.89;1.26) | 1.10<br>(0.86;1.40) | 1.11<br>(0.88;1.41) | 1.12<br>(0.91;1.37) | .. | .. |
|  | ARIC [1, 3] | 2.95*<br>(1.30;6.59) | .. | 2.50*<br>(1.50;4.00) | .. | 1.50*<br>(1.00;2.30) | 1.90*<br>(1.20;3.00) |
|  | CHS [1, 4] | 0.94<br>(0.63;1.41) | .. | .. | .. | .. | .. |
| Arteriovenous<br>nicking | AGES [1, 2] | 1.07<br>(0.93;1.22) | 1.33*<br>(1.09;1.62) | 1.64*<br>(1.36;1.97) | 1.54*<br>(1.31;1.81) | .. | .. |
|  | ARIC [1, 3] | 1.90*<br>(1.25;2.88) | .. | 2.10*<br>(1.40;3.20) | 1.20<br>(0.80;1.60) | 1.20<br>(0.90;1.70) | .. |
|  | CHS [1, 4] | 1.84*<br>(1.23;2.76) | .. | .. | .. | .. | .. |
| Focal narrowing | AGES [1, 2] | 1.08<br>(0.90;1.29) | 1.19<br>(0.92;1.54) | 1.40*<br>(1.09;1.79) | 1.40*<br>(1.12;1.74) | .. | .. |
|  | ARIC [1, 3] | 1.89*<br>(1.22;2.92) | .. | 2.10*<br>(1.40;3.10) | 1.20<br>(0.90;1.70) | 1.10<br>(0.80;1.60) | .. |
|  | CHS [1, 4] | 1.20<br>(0.81;1.78) | .. | .. | .. | .. | .. |
| Arteriolar narrowing | ARIC [1, 2] | 1.74<br>(0.95;3.21) | .. | 1.20<br>(0.80;1.90) | 1.10<br>(0.80;1.50) | 1.00<br>(0.70;1.40) | .. |
|  | CHS [1, 4] | 1.15<br>(0.99;1.26) | .. | .. | .. | .. | .. |
|  | Rotterdam [1, 5] | 1.14<br>(0.91;1.44) | .. | .. | .. | .. | .. |
| Venular dilation | CHS [1, 4] | 1.05<br>(0.93;1.19) | .. | .. | .. | .. | .. |
|  | Rotterdam [1, 5] | 1.07<br>(0.85;1.35) | .. | .. | .. | .. | .. |

Note: Here \* means $p \leq 0.05$ .

**Table 2.** Association of white matter lesion (WML) with the stroke, dementia and deaths based on the outcome of 46 longitudinal studies [11].

| Disease | Hazard Ratio | 95% Confidence Interval |
| --- | --- | --- |
| Stroke | 3.3 | 2.6 to 4.4 |
| Dementia | 1.9 | 1.3 to 2.8 |
| Death | 2.0 | 1.6 to 2.7 |

**Table 3.**
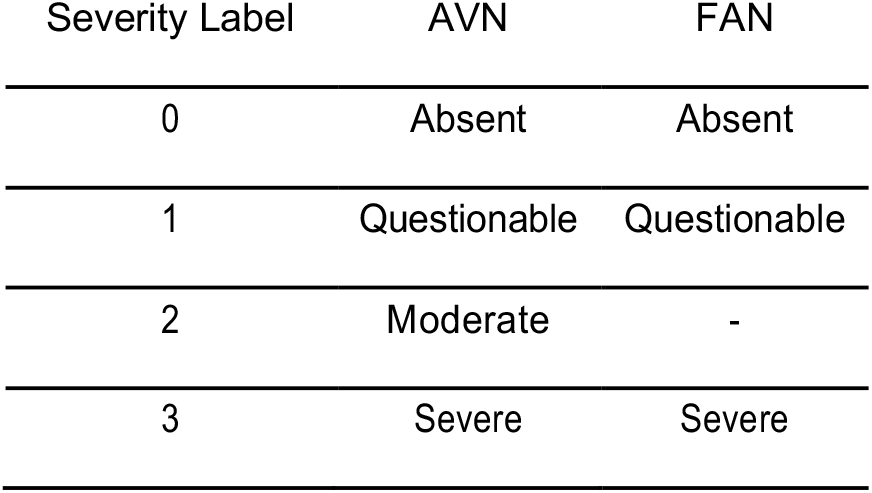
Grading Levels for arteriovenous nicking (AVN) and focal arteriolar narrowing (FAN).

White matter lesions (WMLs) are commonly found on the MRI scans of the brain. The prevalence of WMLs ranges from 11-21% in adults age and around 64% to 94% at the older age of 80 or more [11]. WMLs appear hyper-intensed in the T2 weighted and fluid attenuated inversion recovery (FLAIR) MRI scans as shown in Figure 2. The underlying pathology of these lesions mostly reflects demyelination and axonal loss as a consequence of chronic ischemia caused by cerebral small vessel disease (microangiopathy) [12]. Early diagnosis of WMLs can help the physicians to consider the clinical events associated with it for the preventive treatment. Regular MRI screening of a patient can be done to detect WML in early stage, but it is not practical because of its high expense and inadequacy in the rural areas. Retinal color fundus imaging can be an efficient technology to resolve this problem as it can visualize retinal vascular system non-invasively. Research studies have shown that, retinal microvascular features: AVN and FAN has significant association with WML as mentioned in Table 1. The previous studies establish the association between WML, and retinal microvascular features based on their qualitative measurement. Since manual qualitative assessments are highly subjective and less reproducible. Therefore, in this study, we have analyzed the association between WML, and two prominent retinal vascular features (AVN and FAN) based on their quantified measurements and proposed a retinal vascular features based WML prediction model.

**Figure 2.**
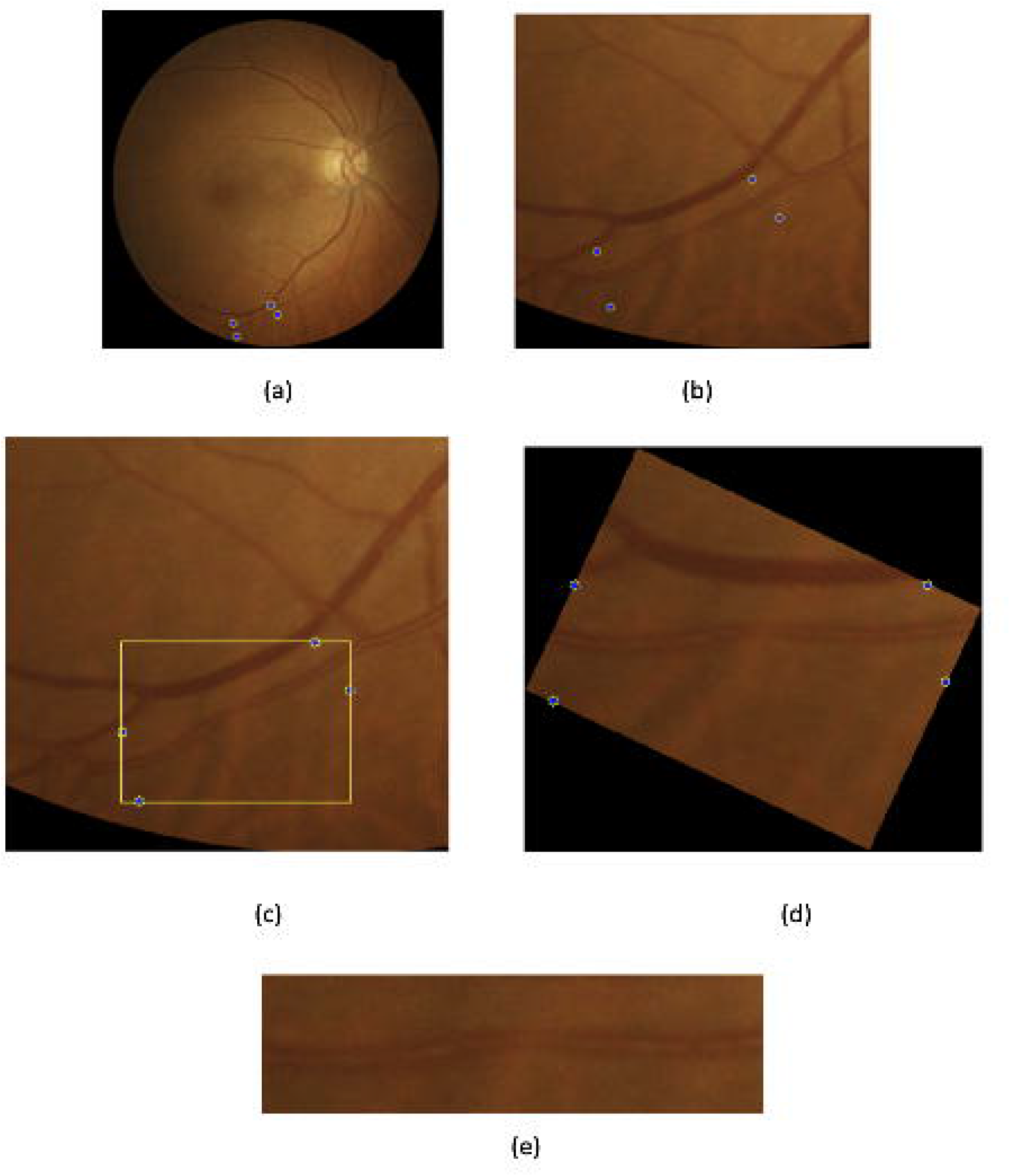
Example of axial fluid attenuated inversion recovery (FLAIR) MRI with (a) minor white matter lesion and (b) extensive WML.

In our proposed model, we use retinal micro-vascular sign to predict the severity of WML load. Focal arteriolar narrowing (FAN) and AVN are two prominent micro-vascular signs in the retina. Arteriovenous nicking (AVN) (as shown in Figure 3) can be defined as the narrowing of venular caliber by a stiff artery at their crossing point and FAN (as shown in Figure 4) is the sudden narrowing of arteriolar width in the retina. FAN and AVN have shown significant association with WMLs load in the brain as mentioned in Table.2. Retinal arteriolar opacity or copper wiring showed association with hypertension which is a risk factor for heart attack and stroke [4, 9, 13]. Therefore, severity of AVN, FAN and Opacification (OP) can be an important parameter for predicting the severity of WML load. In previous studies, manual qualitative grading of AVN and FAN is used to compute their association with WML load. Manual grading can be affected by grader’s fatigue and lack of concentration. As a result, manual grading produces high inter and intra-grader variability and it is highly dependent on the expertise of the grader. Therefore, we hypothesize that computer based quantitative gradings of AVN, FAN and OP can represent their association with WML load more accurately and precisely compared to their manual grading.

**Figure 3.**
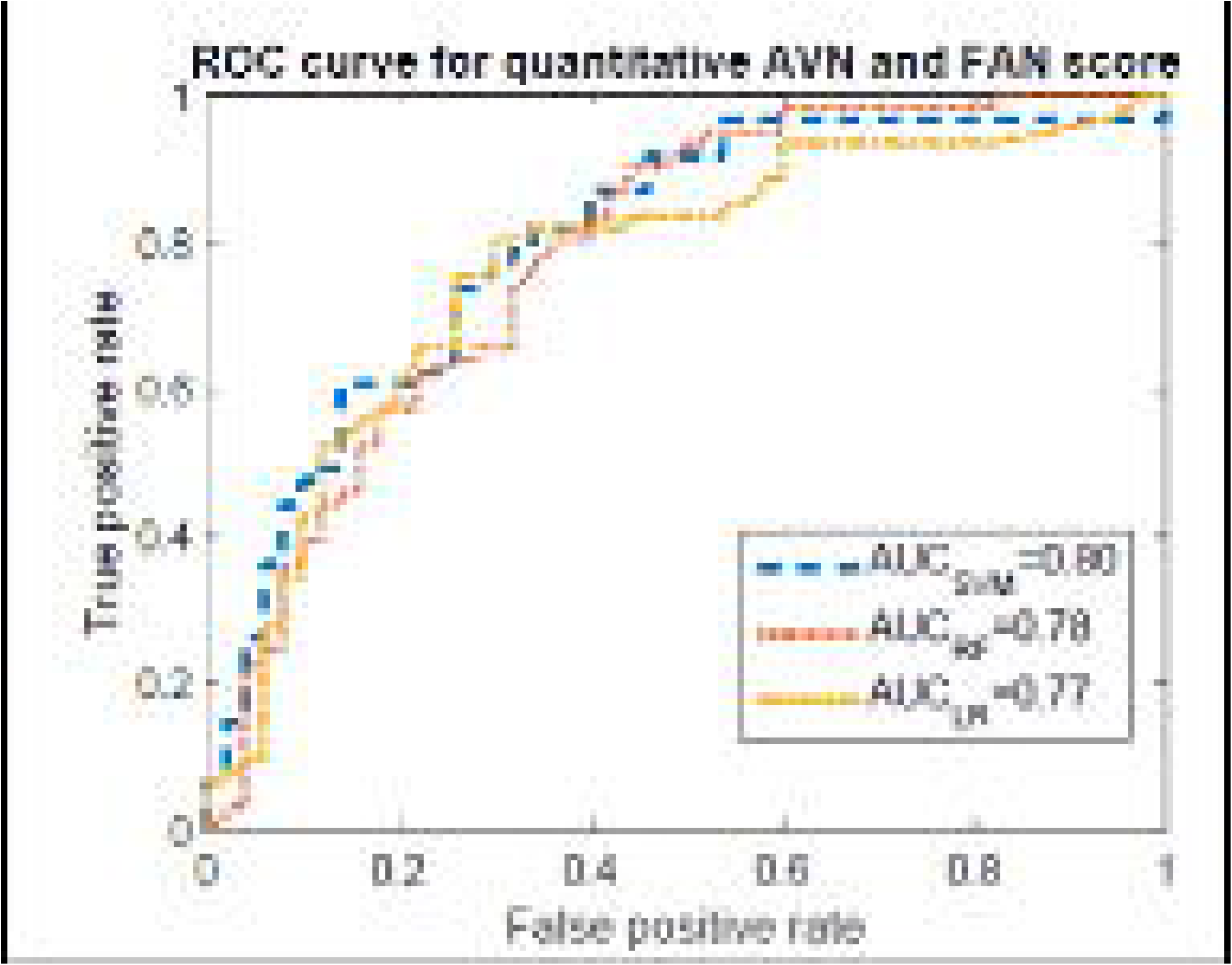
Example of arteriovenus nicking (AVN) affected artery-vein crossover (AV) point (a) and healthy AV crossover point (b).

**Figure 4.**
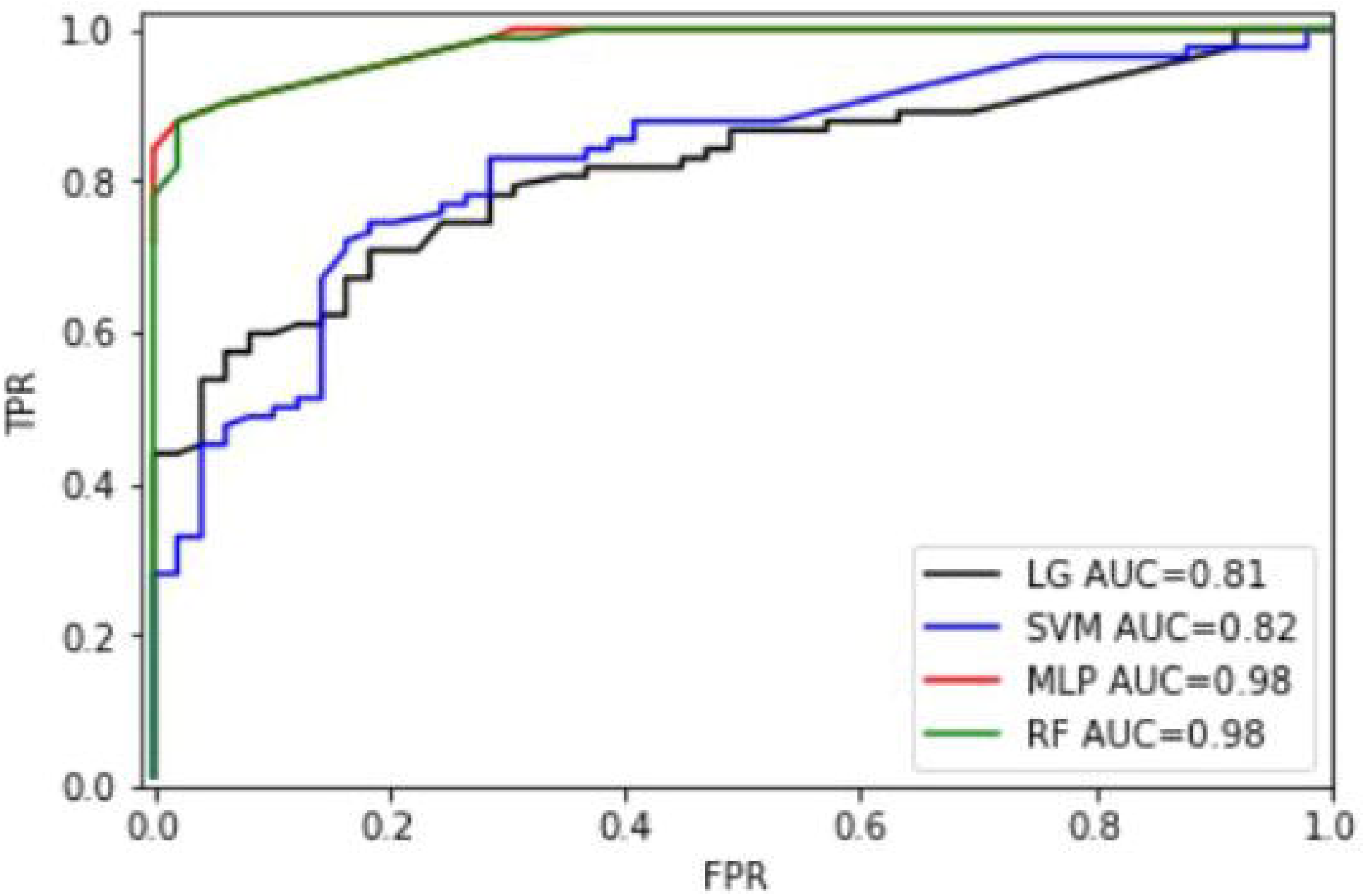
Color retinal image with focal arteriolar narrowing and light reflex (opacification or bright copper wiring) in the center of the artery as sown by arrow.

In this paper, our main contribution is the proposal of a model to estimate and classify the severity of WML load based on the computer aided quantified measurement of AVN, FAN and OP. To best of our knowledge, no previous work has been done on the prediction of WML volume based on the quantified AVN, FAN and OP measurement. In addition, we compare quantitative and qualitative AVN and FAN measurement-based state of the art machine learning prediction algorithms to find out the benefit of the computer aided quantification of AVN and FAN. We evaluate our proposed model on a dataset of 111 patients of ENVISion [14] study. For each of these patients, we have multi-channel MRI images (T1, T2 and FLAIR) and corresponding disc and macula centered retinal images.

The rest of the paper is organized as follows. Section 1 details the methodology used in this study. The experimental results and discussions are presented in section 2, and section 3 concludes the paper.

## 1. Method

The block diagram of the proposed WML volume prediction model is presented in Figure 5. In the proposed WML volume prediction model, quantified retinal micro-vascular signs (AVN,OP and FAN) are extracted as features from a color retinal image. The extracted features are then used to train machine learning models such as *Random Forest, artifical Neural Network, Linear Regression and Support Vector regressor* to estimate the WML volume. The following subsections contains the description of our dataset, feature extraction step and training machine learning models: Random Forest, artifical Neural Network,, *Linear Regression* and support vector for WML volume prediction.

**Figure 5.**
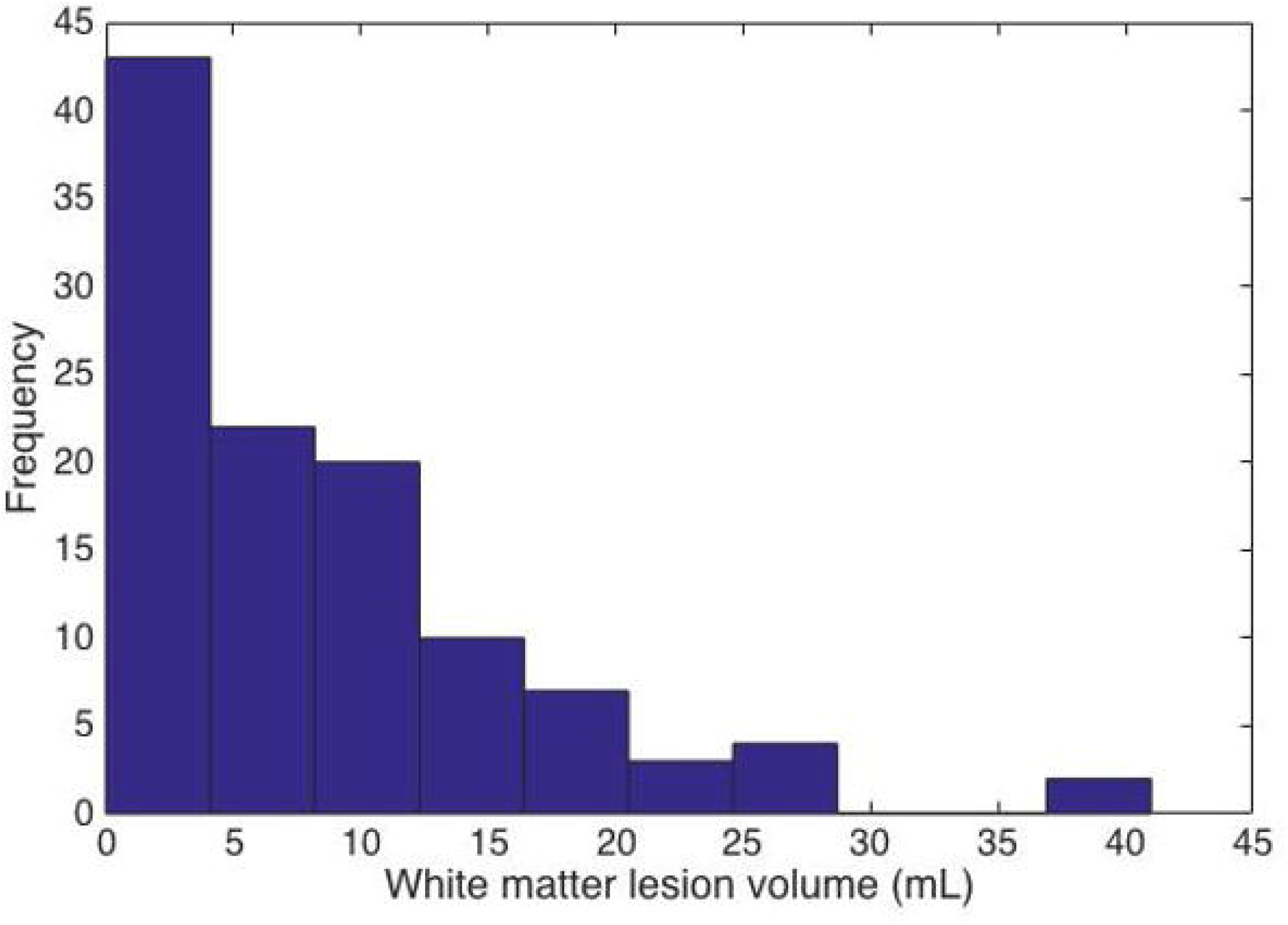
Block diagram of the proposed white matter lesion (WML) volume prediction model.

### 1.1 Dataset

Brain MRI and retinal vascular imaging data used in this work are obtained from the ENVISion [14] study. All participants have given written consent separately for the ENVIS-ion trial. Protocols of ENVIS-ion study have been approved by the Monash University health and research ethics committee. Our study is approved as a low-risk project under the ENVIS-ion study by the Alfred hospital ethics committee, Melbourne, Victoria. Therefore, we have access to only imaging data of this study.

A 111 standardized Brain MRI sequences are acquired to allow morphologic, microstructure and functional assessment. First, a volumetric 3D T1-weighted sequence is used to obtain high resolution anatomical information. The size of each subject’s voxel set is 166×256×256 and the resolution is 1:2× 0:94× 0:94 mm3. This is followed by the FLAIR and T2 sequences used to assess inflammation and signal alteration in brain tissues. For them size of each subject’s voxel set is 256×256×36 and resolution is 0:94 0:94 4 mm3.

On the other-hand, two color retinal photographs, centered on the optic disc and macula, respectively, is taken from both left and right eye of the participant. The photographs are taken after 5 minutes of dark adaption, without pharmacological pupil dilation, using a Canon NMR 45 digital fundus camera. The resolution of retinal images is 4752 pixelsx3168 pixels.

The manual segmentation of WML is time consuming therefore, we use an automated method proposed in [16] to segment WML. In [15] a wide range of features are used which contain multi-channel local intensity, textural and spatial information to train a Random Forest (RF) classifier to compute the posterior probability of a voxel as a lesion. Then the RF computed lesion probability of each voxel is incorporated into a Markov Random Field (MRF) to obtain the final segmentation of white matter lesion. After the automatic segmentation of white matter lesions, the false positive and missing lesions are manually corrected by an expert grader using ITK-SNAP [16]. After the manual correction of WML, their volume is computed by using the voxel’s resolution and size.

### 1.2 Feature extraction

As mentioned in section 1.1, our dataset contains retinal images and MRI images of 111 patients. For each of these patients, AVN is graded into four levels (0=absent, 1=questionable, 2=moderate and 3=severe) and FAN is graded into 3 levels (0=absent, 1=questionable and 3=severe) by two expert graders of the Centre for Eye Research Australia (CERA). The manually graded AVN and FAN segments are then quantified by two semi-automatic methods proposed in [17] and [18].

We have created the feature set using the quantified measurement of AVN and FAN. The details of the feature extraction step are given in the following subsections.

#### 1.2.1 Quantified score of arteriovenous nicking (AVN)

We compute the amount of narrowing in the vein near the artery vein (AV) crossover point to quantify the severity of AVN using the method proposed in [18]. The procedures of retinal vessel segmentation, AV crossover point detection, region of interest selection, artery vein classification, separation of venular segment, vein edge and centerline refinement and width computation are explained in [18] (as shown in Figure 6). We quantify the AVN by using the characteristics of the vein widths as shown in Figure. 7. Suppose that the vein segment 1 in Figure 7 is made up of n vessel cross-sectional (each white line) widths, *w* = {*w*_*i*_ | *i* ∈1,…, *n*}(the vessel widths *w*_*i*_ are indexed and ordered so that w1 is the width closest to the crossover point, while *w*_*n*_ is the furthest). W is approximated as the median value of its measurements, or W = median(w). Suppose that N is the number of successive cross-sections, starting from the first measurement w1, with the measurements lower than normal vessel width W, *WC* is computed as the minimum value of the first N vessel widths, or *W*_*C*_ = min{*w*_*i*_|*i* ∈1,…, *N*}. Then the AVN score for vein segment 1 is computed by the following equation:

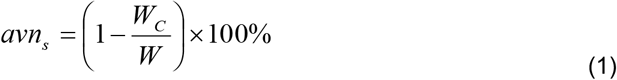

**Figure 6.**
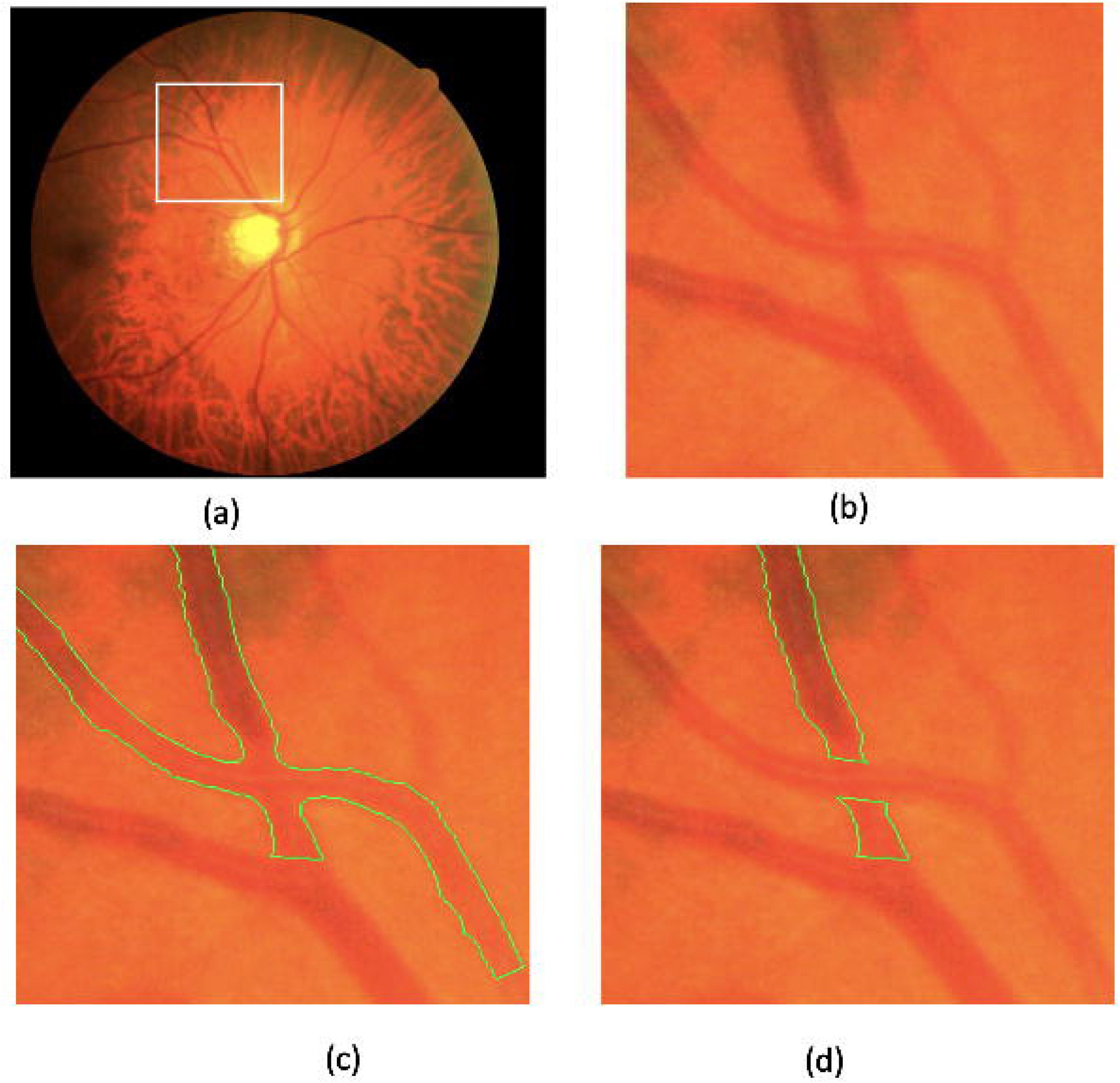
Output images of the different steps of image processing module for arteriovenous nicking (AVN) quantification (a) original image; (b) region of interest; (c) simplified segmentation of the artery vein crossover point; (d) detection of venular segments.

**Figure 7.**
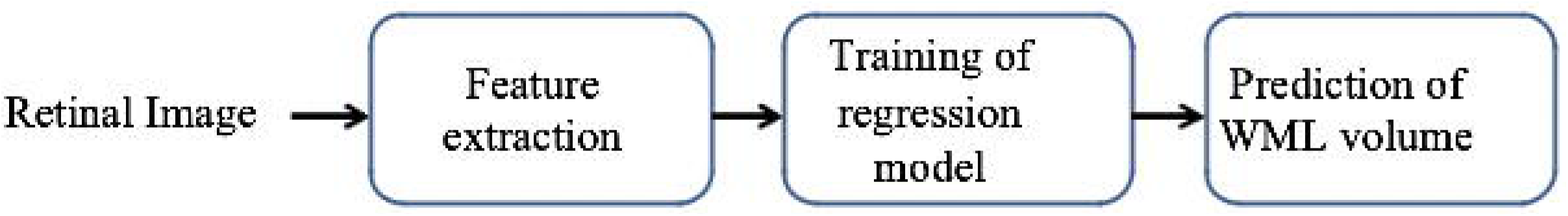
Automatically computed width for vein segment 1 and vein segment 2.

Since each crossover point has two venular segment, therefore, we will get two avn_s_ score such as *avn*_*s1*_ for vein segment 1 and *avn*_*s2*_ for vein segment 2. The max (avn_s1_, avn_s2_) is chosen as the final avn score for the respective AV crossover point.

We select maximum of the AVN scores because it has shown highest correlation with the corresponding manual grading of AVN severity compared to the summation and average of the AVN scores of two vein segments correspond to an AV cross-over point.

#### 1.2.2 Quantified score of focal arteriolar narrowing (FAN)

We adopted a semi-automatic approach proposed in [18] to quantify the FAN. The region of interest is selected by clicking four points around the vessel as shown in Figure 8. Then we use the method proposed in [17] to crop the selected region in such a way that the vessel will be always located at the center of the ROI. Following that the cropped image is affinely transformed as mentioned in [18] to horizontally align the vessel position with the center of the image as shown in Fig. 8(b)

**Figure 8.**
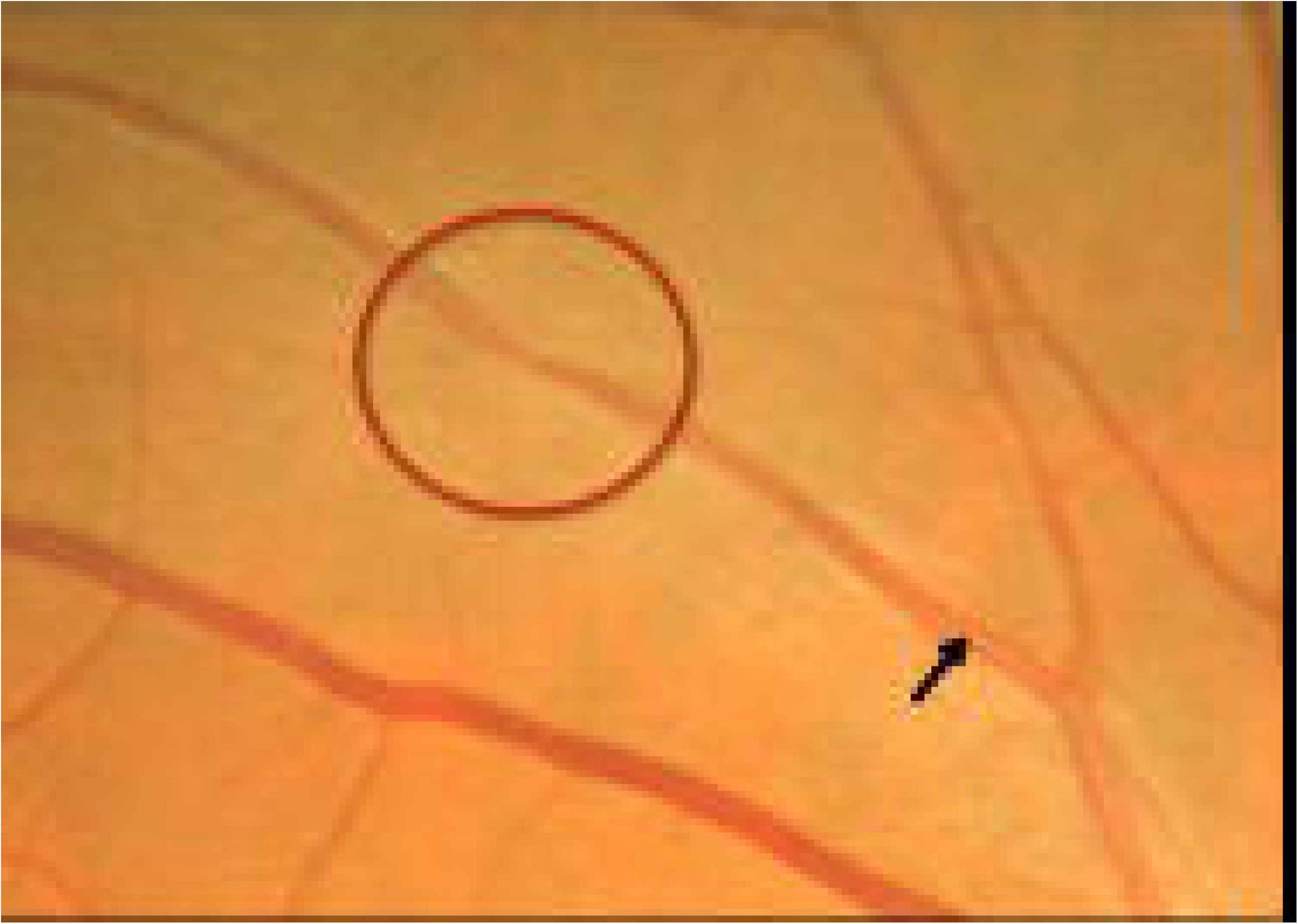
(a) Original retinal image with user selected region of interest points marked by yellow cross point, (b) zoomed original image, (c) cropped region of interest (marked by yellow box),(d) affine translation of the region of interest and (e) segmented vessel region.

This spatial normalization of the vessel makes the mapping of edges easier. We apply an edge preserving smoothing filter proposed in [19] to suppress the noise (Fig. 8(c)). After smoothing the vessel region, a Canny edge detector [20] is applied in the spatially normalized ROI to detect the edges (Fig. 8(d)). Vessel edges are mapped from the detected edges (Fig. 8(e)) by using Dijkstra’s shortest path algorithm [21]. After identifying vessel edges, its width (as shown in Fig. 8(g)) is measured through mapping edge pixel pairs from both edges as proposed in [17].

For each arteriolar segment, we compute fan_s_ from the vessel widths of that segment to measure the amount of narrowing. Suppose that the segment is made up of n arteriolar width measurements as shown in Figure 9, *w* = {*w*_*i*_|*i* ∈1,…, *n*}. The average of the width measurements is computed as follows:

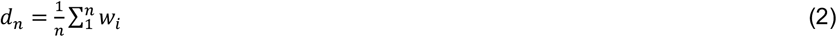

**Figure 9.**
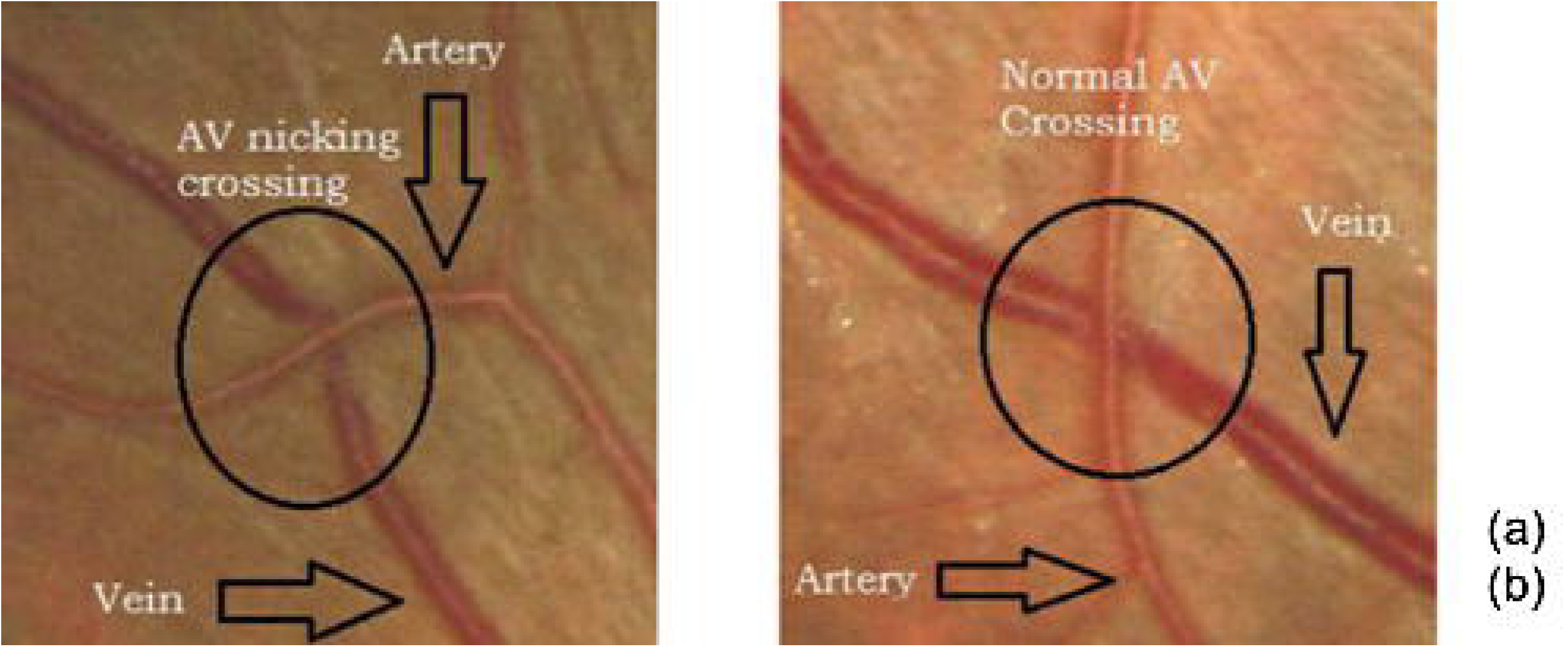
(a) Vessel segment marked with width measurements, (b) plot of the cross-sectional widths of the measurement.

Let j is the index of the minimum width measurement 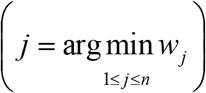 then the *j* th width measurement is smoothed by using a boxfilter of size n as follows:

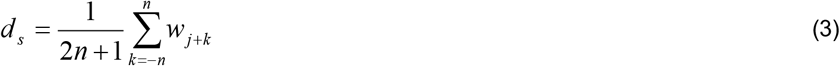

Finally, the narrowing in percentage is computed as follows:

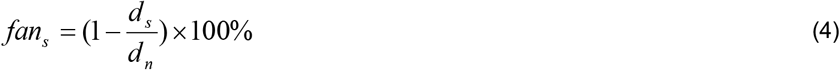

For each subject, we compute the number of AVN and FAN cases and statistical information (mean, median, standard deviation, summation) of the corresponding quantified AVN and FAN scores (*avn*_*s*_ and *fan*_*s*_) as shown in Table 4. Illustration of the feature set construction for a patient is shown in Figure. 10. Here, patient-1 has 7 AVN and 5 FAN affected vessels.

**Table 4.** Description of the feature set.

| Name | Description |
| --- | --- |
| #AVN | #AVN is the number of AVN cases |
| #FAN | #FAN is the number of FAN cases |
| #OP | #OP is the number of OP cases |
| maxAVN/ maxFAN / maxOP | Maximum of the quantified AVN / FAN / OP scores |
| meanAVN/ meanFAN / meanOP | Mean of the quantified AVN / FAN / OP scores |
| medianAVN/ medianFAN / medianOP | Median of the quantified AVN / FAN / OP scores |
| sumAVN/ sumFAN / sumOP | Summation of the quantified AVN / FAN / OP scores |
| stdAVN/ stdFAN / stdOP | Standard deviation of the quantified AVN / FAN / OP scores |

**Figure 10.**
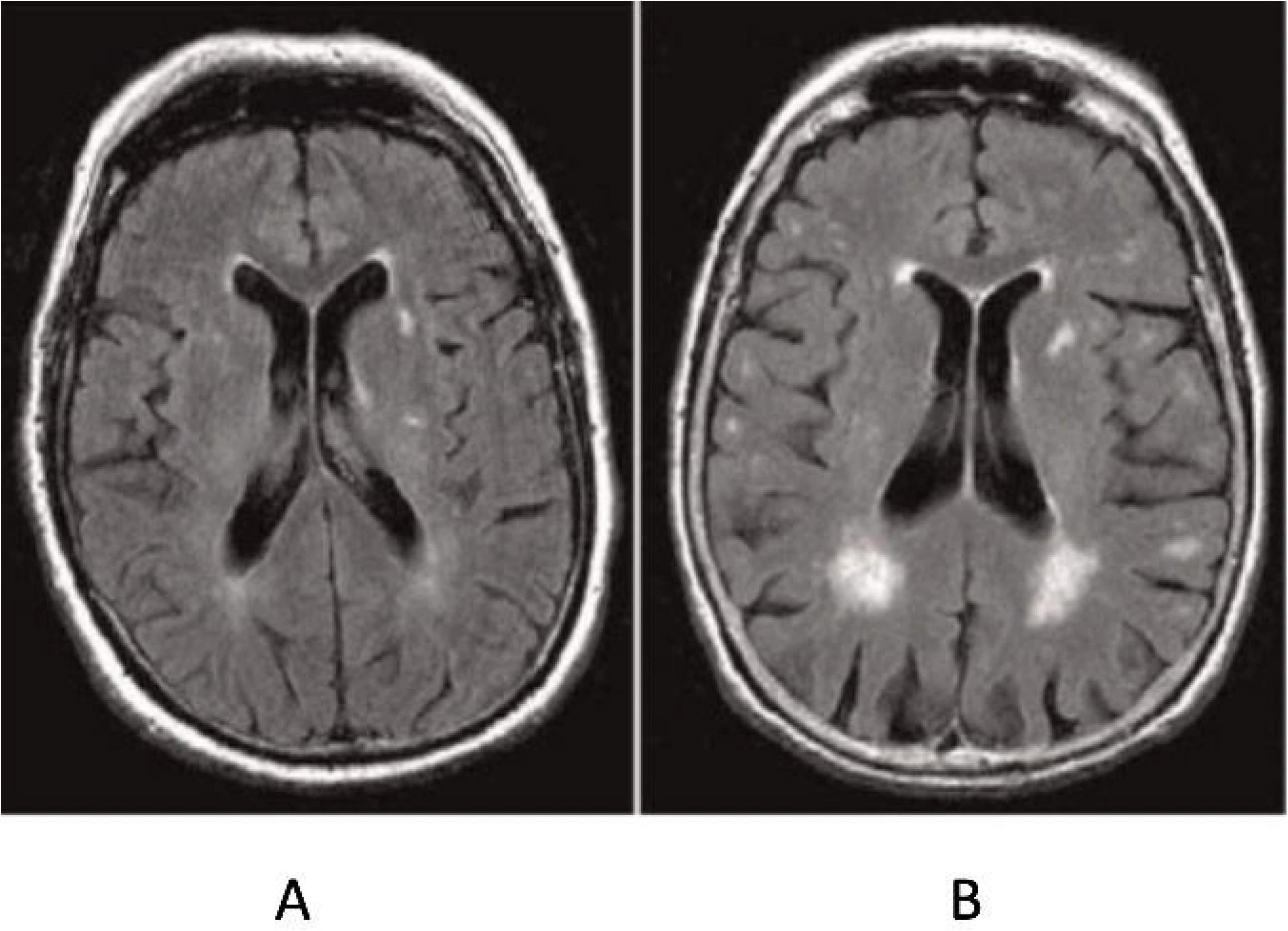
Distribution of WML over the study sample

**Figure 11.**
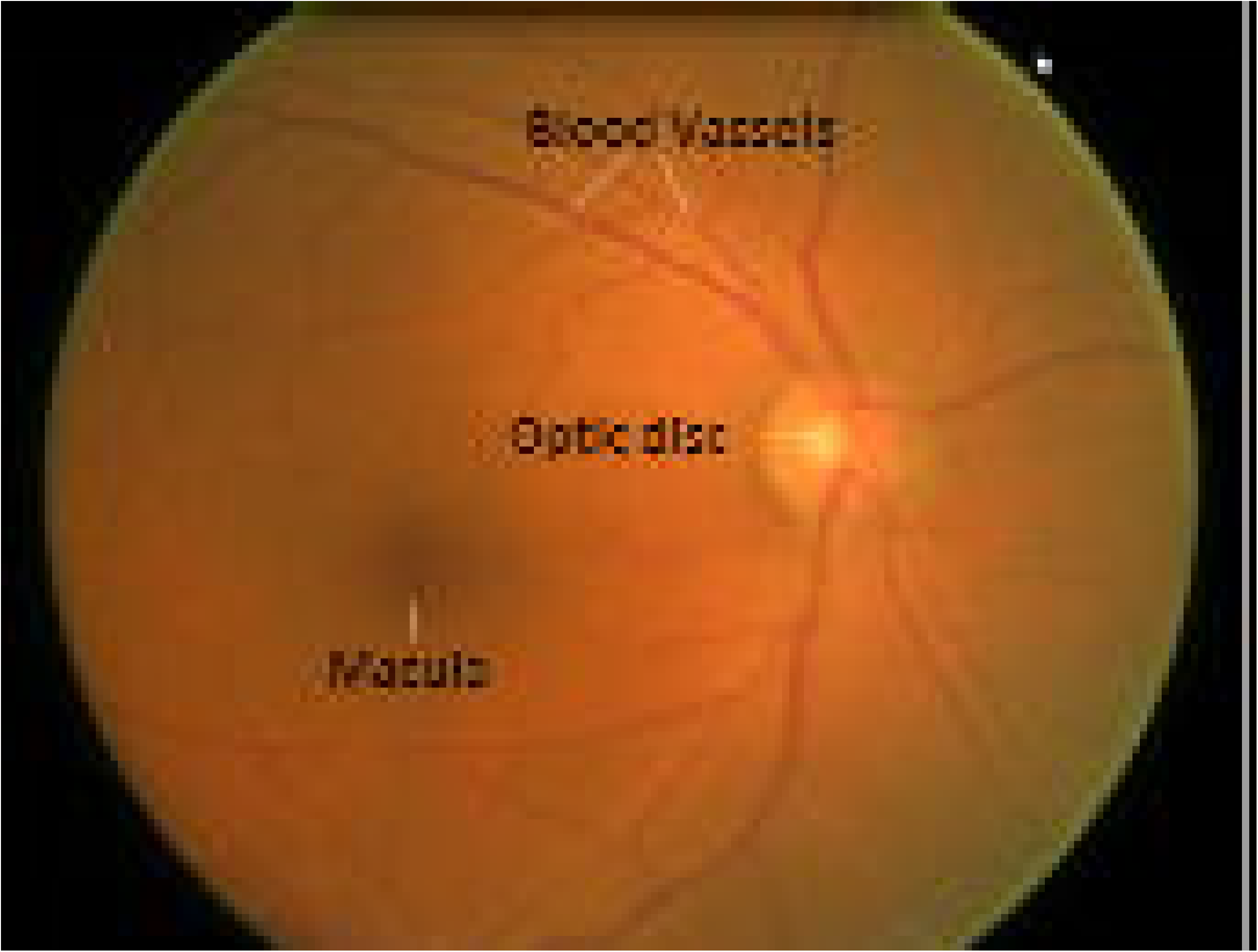
ROC curve for quantitative feature based artifical neural network (ANN), support vector machine (SVM), random forest, (RF), and linear regression (LR) classifiers.

**Fig. 12.** ROC curve for SVM random forest (RF) and linear regression (LR) classifiers (top 3 classifiers).

### 1.3 Prediction and classification of white matter lesion volume

We have compared the state of the art machine learning algorithms to obtain the optimal WML prediction model using the feature set proposed in Table 4. A brief description of these models are given in the following sub sections.

#### 1.3.1 Support Vector machine (SVM)

SVM [25] is a learning system that uses higher dimensional feature space. A support vector machine constructs a hyperplane in a higher dimensional space which can be used for classification or regression. Intuitively, a good separation is achieved by the hyperplane that has the largest distance to the nearest training data point of any class (so-called functional margin), since in general the larger the margin the lower the generalization error of the classifier. In our implementation, we used the kernel radial basis function (rbf), with a C value of 1.0, and a degree 3.

#### 1.3.2 Random Forest (RF)

A Random Forest [22] consists of a various number of individual decision trees. It is an ensemble estimator which gives lower variance. We used the scikit-learn implementation of random forest machine learning model. The most important parameter in this case is the number of estimators i.e., the number of individual decision trees. We experimented with different number of decision trees ranging from 10 to 600. We got the best accuracy when we used 400 trees. All other parameters such as criterion and max_depth were set as default value of scikit-learn implementation. We used 400 trees, allowed nodes to expand until all leaves are pure, minimum samples split of 2, and minimum samples required to be at leaf node to be 1.

#### 1.3.3 Artifical Neural Network (ANN)

Artifical Neural Network (ANN) [22] is a supervised machine learning algorithm that has the capability to learn non-linear models. It can have one or more non-linear layers in between the input layer and the output layer. These non-linear layers are called hidden layers. Our ANN has 3 hidden layers each containing of 100 hidden units called neurons. We used the scikit-learn implementation of artifical neural network. We used ‘lbfgs’ solver for weight optimization as our dataset was small. Because in case of small datasets, ‘lbfgs’ converges faster and performs better [22]. Rectified Linear Unit (ReLU) was used as activation function. All other parameters were set as default.

#### 1.3.4 Linear Regression (LR)

Linear Regression is an approach to model the relationship between two variables, dependent and independent variables, by fitting a linear equation to the data. The relationships are modeled using linear predictor functions whose parameters are estimated using the observed data. We used the scikit-learn implementation of linear regression in our study. The regressors would be normalized before regression by subtracting the mean and dividing by the “l2-norm”.

### 1.4 Evaluation

We use mean square error (MSE) as shown in equation 6 to compute the accuracy of the prediction of WML volume. Here, *Ŷ*_*i*_ represents the predicted volume and *Y*_*i*_

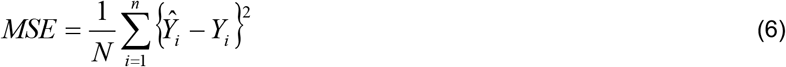

We select a cut-off value of 5mL to classify the severity of WML volume. The severity of the WML load is usually classified into four levels [23] as shown in Table 5. In our data set, the number of subjects is comparatively small in severity level 3 and 4, therefore, we combine severity level 2, 3 and 4 into one class named as present and severity level 1 is considered as absent class. We apply the same cut-off value (5 mL) in the predicted WML volume to classify them into absent and present class. The distribution of WML over the study sample is presented in Figure 10. Following that we compute sensitivity (SEN), specificity (SPE), positive predictive value (PPV), negative predictive value (NPV) and

**Table 5.** Classification of the severity of white matter lesion volume [23]

| Severity Level | Lesion volume (LV) | Subject | Class |
| --- | --- | --- | --- |
| 1 | LV < 5 mL | 52 | absent |
| 2 | 5 mL < LV < 10 mL | 31 |  |
| 3 | 10 mL < LV < 15 mL | 12 | Present |
| 4 | LV > 15 mL | 16 |  |

*F*_1_ score to evaluate the classification accuracy. All these measurement metrics are computed by using TP (true positive), FP (false positive), TN (true negative) and FN (false negative) as shown in the following equations:

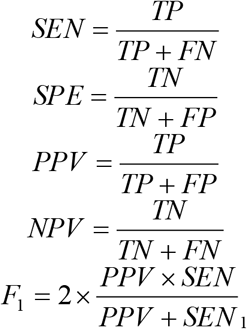

As mentioned earlier, we use ten-fold cross validation to evaluate our prediction of WML volume. To remove the training data bias, we shuffle the subjects randomly to produce ten different permutations of the subjects. For each random permutation, we run ten-fold cross validation resulting 100 evaluations. The mean MSE, SEN, SPE, PPV, NPV and F_1_ score are reported in the result section.

## 2. Results and Discussion

Table 6 and Table 7 shows the prediction accuracy of the proposed method for different input parameter sets. We use four state of the art classifier models: random forest (RF) classifier, artifical neural network (ANN) classifier, support vector machine (SVM) classifier and logistic regression (LR) classifier to predict the WML volume absent or present classification. Table 6 and Table7 show that, all of these models (SVM, LR, ANN and RF) are showing better prediction accuracy for combination of all three of quantitative AVN, FAN OP scores compared to combination of any two features/input parameters of AVN, OP and FAN scores or using AVN, OP and FAN individually. We apply a cut-off value of 5 mL as mentioned in section 1.4 to classify the actual and predicted WML volume into absent and present class and compute the classification accuracy. Among the four models, quantitative AVN, OP and FAN score-based RF classifier is showing the highest sensitivity of 0.84 with a PPV score of 0.96. In contrast to that, the maximum specificity of 0.99 is shown by quantitative AVN, OP and FAN score-based ANN with a negative predictive value of 0.88. The overall classification accuracy is computed by F1 score, where quantitative AVN, OP and FAN score-based RF and ANN classifier both are showing highest F1 score of 0.92. From Table 6 and Table 7 we can draw a conclusion that proposed quantitative AVN, OP and FAN score-based models are showing better prediction accuracy compared to the qualitative AVN and FAN score-based models.

**Table 6.** Prediction and classification accuracy of artificial neural network (multi-layer perceptron), support vector classifier, logistic regression classifier and random forest classifier using combination of quantitative arteriovenular narrowing (AVN), Opacity (OP) and focal narrowing scores(FAN)as Input Parameter.

| Classifier | Input Parameter | Accuracy ( % ) | SEN | SPE | PPV | NPV | F1 |
| --- | --- | --- | --- | --- | --- | --- | --- |
| RF | AVN,FAN & OP | 91.6 (85.5 to 95.7)% | 0.84 | 0.97 | 0.96 | 0.89 | 0.92 |
| ANN | AVN,FAN & OP | 91.6 (85.5 to 95.7)% | 0.83 | 0.99 | 0.98 | 0.88 | 0.92 |
| SVM | AVN,FAN & OP | 77.1 (69.0 to 84.0)% | 0.68 | 0.83 | 0.73 | 0.79 | 0.77 |
| LR | AVN,FAN & OP | 71.8 (63.2 to 79.3)% | 0.65 | 0.75 | 0.53 | 0.83 | 0.71 |

**Table 7.** Prediction and classification accuracy of Artificial Neural Network (ANN) ,support vector classifier, logistic regression classifier and random forest classifier using combinations of 2 features among quantitative arteriovenular narrowing (AVN),opacity (OP) and focal narrowing scores(FAN) as Input Parameter at a time and using AVN, OP and FAN individually as Input Parameter.

| Classifier | Input Parameter | Accuracy ( % ) | SEN | SPE | PPV | NPV | F1 |
| --- | --- | --- | --- | --- | --- | --- | --- |
| RF | AVN, FAN | 88.5 (81.8 to 93.5)% | 0.80 | 0.95 | 0.92 | 0.87 | 0.89 |
|  | AVN, OP | 85.5 (78.3 to 91.0)% | 0.81 | 0.88 | 0.80 | 0.89 | 0.85 |
|  | FAN,OP | 79.4 (71.5 to 86.0)% | 0.66 | 0.94 | 0.92 | 0.72 | 0.79 |
| ANN | AVN, FAN | 88.5 (81.8 to 93.5)% | 0.79 | 0.96 | 0.94 | 0.85 | 0.89 |
|  | AVN, OP | 85.5 (78.3 to 91.0)% | 0.81 | 0.88 | 0.80 | 0.89 | 0.85 |
|  | FAN,OP | 79.4 (71.5 to 86.0)% | 0.67 | 0.92 | 0.90 | 0.73 | 0.80 |
| SVM | AVN, FAN | 74.1 (65.7 to 81.3)% | 0.66 | 0.79 | 0.64 | 0.80 | 0.74 |
|  | AVN, OP | 68.0 (59.2 to 75.8)% | 0.58 | 0.74 | 0.55 | 0.76 | 0.68 |
|  | FAN,OP | 66.4 (57.6 to 74.4)% | 0.56 | 0.72 | 0.49 | 0.77 | 0.66 |
| LR | AVN, FAN | 74.8 (66.5 to 82.0)% | 0.69 | 0.78 | 0.59 | 0.84 | 0.74 |
|  | AVN, OP | 65.7 (56.9 to 73.7)% | 0.56 | 0.69 | 0.39 | 0.82 | 0.64 |
|  | FAN,OP | 66.4 (57.6 to 74.4)% | 0.55 | 0.73 | 0.53 | 0.74 | 0.66 |
| RF | AVN | 72.5 (64.0 to 80.0)% | 0.60 | 0.85 | 0.80 | 0.68 | 0.73 |
|  | FAN | 76.3 (68.1 to 83.3)% | 0.64 | 0.89 | 0.86 | 0.71 | 0.77 |
|  | OP | 75.5 (67.3 to 82.6)% | 0.90 | 0.73 | 0.39 | 0.98 | 0.73 |
| ANN | AVN | 72.5 (64.0 to 80.0)% | 0.60 | 0.85 | 0.80 | 0.68 | 0.73 |
|  | FAN | 76.3 (68.1 to 83.3)% | 0.63 | 0.90 | 0.88 | 0.70 | 0.77 |
|  | OP | 75.6 (67.3 to 82.7)% | 0.90 | 0.73 | 0.39 | 0.98 | 0.72 |
| SVM | AVN | 67.9 (59.2 to 75.8)% | 0.56 | 0.76 | 0.63 | 0.71 | 0.68 |
|  | FAN | 63.4 (54.5 to 71.6)% | 0.67 | 0.63 | 0.04 | 0.99 | 0.51 |
|  | OP | 62.6 (53.7 to 70.9)% | undefined | 0.63 | 0.0 | 1.0 | 0.48 |
| LR | AVN | 66.4 (57.6 to 74.4)% | 0.55 | 0.73 | 0.55 | 0.73 | 0.66 |
|  | FAN | 64.1 (55.3 to 72.3)% | 0.67 | 0.64 | 0.08 | 0.98 | 0.54 |
|  | OP | 64.1 (55.3 to 72.3)% | 1.00 | 0.64 | 0.04 | 1.00 | 0.52 |

Finally, it can be noted that quantified AVN, FAN and OP scores are showing better accuracy in predicting the WML volume class compared to the corresponding qualitative grading, which confirms a number of research studies [24-26] where quantitative grading shows better accuracy than qualitative grading in disease prediction. Since the retinal imaging technology is a cheaper option, and it can be installed in low socio-economic counties and rural areas, a retinal image based accurate white matter prediction model can be helpful for clinicians to identify those patients who need further detailed diagnosis. Our future goal is to evaluate our model on a large scale and longitudinal dataset.

We have shown that we can predict WML with higher accuracy using quantified retinal FAN and AVN than the qualitative FAN and AVN. We compute the features to train the machine-learning classifier (i.e., SVM) to predict the brain WML severity by WML volume. WML severity levels are classified as: none (<5ml), mild (>5ml and <10ml), moderate (>10ml and <15ml), and severe (>15ml). We evaluate our proposed model on a dataset of 111 patients taken from the ENVIS-ion study which has retinal and MRI images for each patient. Our model shows high accuracy in estimating the WML volume/severity (none, mild, moderate, and severe); the mean square error (MSE) between our predicted WML load and manually annotated WML load is 0.13. Proposed WML prediction model shows high classification accuracy with **AUC 0.80 (the expert graded i.e**., **qualitative AVN and FAN score based WML prediction AUC 0.69)** in classifying the healthy patients having mild and severe WML load. The ROC in Fig. 13 shows that we can screen more than 90% of individuals with WML while picking the 50% normal individuals at high risk. We note that **we only considered two features** and did not consider the proposed parameters (age, gender, etc. which were not provided yet).

## 3. Conclusion

In this paper, a quantified AVN, FAN and OP score-based prediction model is proposed to predict the severity of WML volume in the brain. The prediction model is evaluated on a dataset of 111 patients of ENVISion study which has MRI and retinal image for each subject. Quantification of WML volume is done by using our proposed automatic software, which is further refined by manual correction. Arteriovenous nicking (AVN) and focal arteriolar narrowing (FAN) are graded by the expert graders of Center for Eye Research Australia. Then each manually graded AVN and FAN affected vessel segment is quantified by using our proposed computer aided diagnostic (CAD) methods. We adopted four state of the art classifiers to classify the severity of WML volume. Proposed quantified AVN, OP and FAN scores significantly improve the classification accuracy compared to the expert graded qualitative AVN, OP and FAN scores for each of these classifiers. We obtain the optimal classification accuracy (*F*_1_**score = 0.92)** by using **both RF and ANN** classifier. Our future goal is to include patients meta data information (age, sex, gender, ethnic origin, etc.) into our model and evaluate the performance on a large-scale and longitudinal dataset.

Proposed WML prediction model shows high classification accuracy with **AUC 0.80 (compared to the expert graded i.e**., **qualitative AVN and FAN score based WML prediction, AUC 0.69)** in classifying the healthy patients having mild and severe WML load.

WML may be an indicator for stroke and Alzheimer’s disease prediction, thus our model can help screening for early detection of indicuviduals at risk of stroke and Alzheimer’s disease. At present, there is affordable little opportunity for treatment of Alzheimer’s outside of palliative care. Recent clinical findings suggest that successful treatment needs to start in the prodromal stages of the disease[5]. Doctors can’t definitely diagnose AD until after death when they can closely examine the brain under a microscope [6]. However, regular Magnetic Resonance Imaging (MRI) of the brain can be used to find out symptoms of AD [7], but regular MRI is impractical for the screening of a patient because of its high cost and inaccessibility in remote areas. Therefore, retinal imaging can be a key for screening in the primary care level. Similarly, we can utilize the screening tool for identification of individuals at risk of incident stroke.

## Data Availability

N/A

## References

[1] C. Wilkinson et al., “Proposed international clinical diabetic retinopathy and diabetic macular edema disease severity scales,” Ophthalmology, vol. 110, no. 9, pp. 1677–1682, 2003.

[2] T. Wong and P. Mitchell, “Hypertensive retinopathy,” N Engl J Med, vol. 351(22), pp. 2310–7, 2004.

[3] P.M. D’onofrio and P. D. Koeberle, “What can we learn about stroke from retinal ischemia models?,” Acta Pharmacologica Sinica, vol. 34, no. 1, pp. 91–103, 2013.

[4] S. M. Heringa, W. H. Bouvy, E. Van Den Berg, A. C. Moll, L. J. Kappelle, and G. J. Biessels, “Associations between retinal microvascular changes and dementia, cognitive functioning, and brain imaging abnormalities: a systematic review,” Journal of Cerebral Blood Flow & Metabolism, vol. 33, no. 7, pp. 983–995, 2013.

[5] T. Y. Wong and R. McIntosh, “Systemic associations of retinal microvascular signs: a review of recent population-based studies,” Ophthalmic and Physiological Optics, vol. 25, no. 3, pp. 195–204, 2005.

[6] A. R. Sharrett, “A review of population-based retinal studies of the microvascular contribution to cerebrovascular diseases,” Ophthalmic epidemiology, vol. 14, no. 4, pp. 238–242, 2007.

[7] A. Bhuiyan, R. Kawasaki, E. Lamoureux, T. Y. Wong, and K. Ramamohanarao, “Retinal Vascular Features for Cardio Vascular Disease Prediction: Review,” Recent Patents on Computer Science, vol. 3(3), pp. 164–175, 2010.

[8] A. C. Qiu et al., “Cerebral microbleeds, retinopathy, and dementia the ages-reykjavik study,” Neurology, vol. 75 (24), pp. 2221–2228, 2010.

[9] L. S. Cooper, “Retinal microvascular abnormalities and MRI-defined subclinical cerebral infarction - The atherosclerosis risk in communities study,” (in English), Stroke, vol. 37, no. 1, pp. 82–86, 2006.

[10] M. L. Baker et al., “Retinal microvascular signs, cognitive function, and dementia in older persons the cardiovascular health study,” Stroke, vol. 38(7), pp. 2041–2047, 2007.

[11] S. Debette and H. Markus, “The clinical importance of white matter hyperintensities on brain magnetic resonance imaging: systematic review and meta-analysis,” BMJ, vol. 341:c3666, 2010.

[12] M. Mortamais et al., “Spatial distribution of cerebral white matter lesions predicts progression to mild cognitive impairment and dementia,” PloS one, vol. 8, no. 2, p. e56972, 2013.

[13] C. Qiu et al., “Cerebral microbleeds, retinopathy, and dementia the ages-reykjavikstudy,” Neurology, vol. 75(24) pp. 2221–2228, 2010.

[14] C. M. Reid et al., “Aspirin for the prevention of cognitive decline in the elderly: rationale and design of a neuro-vascular imaging study (ENVIS-ion),” BMC Neurology, vol. 12:3, pp. 1–9, 2012.

[15] P. K. Roy et al., “Automated segmentation of white matter lesions using global neighbourhood given contrast feature-based random forest and Markov random field,” in 2014 IEEE International Conference on Healthcare Informatics, 2014: IEEE, pp. 1–6.

[16] P. A. Yushkevich et al., “User-guided 3D active contour segmentation of anatomical structures: significantly improved efficiency and reliability,” Neuroimage, vol. 31, no. 3, pp. 1116–1128, 2006.

[17] P. K. Roy, A. Hussain, A. Bhuiyan, R. Kawasaki, and K. Ramamohanarao, “A robust and reliable quantification method for Focal Arteriolar Narrowing in color retinal image,” in 2015 IEEE 12th International Symposium on Biomedical Imaging (ISBI), 2015: IEEE, pp. 1510–1513.

[18] P. K. Roy, U. T. Nguyen, A. Bhuiyan, and K. Ramamohanarao, “An effective automated system for grading severity of retinal arteriovenous nicking in colour retinal images,” in 2014 36th Annual International Conference of the IEEE Engineering in Medicine and Biology Society, 2014: IEEE, pp. 6324–6327.

[19] A. Bhuiyan et al., “Retinal artery and venular caliber grading: A semi-automated evaluation tool,” Computers in biology and medicine, vol. 44, pp. 1–9, 2014.

[20] K. He, J. Sun, and X. Tang, “Guided image filtering,” in European conference on computer vision, 2010: Springer, pp. 1–14.

[21] J. Canny, “A computational approach to edge detection,” IEEE Transactions on pattern analysis and machine intelligence, no. 6, pp. 679–698, 1986.

[22] C. Shackelford, G. Long, J. Wolf, C. Okerberg, and R. Herbert, “Qualitative and quantitative analysis of nonneoplastic lesions in toxicology studies,” Toxicologic pathology, vol. 30, no. 1, pp. 93–96, 2002.

[23] C.-C. Chang and C.-J. Lin, “LIBSVM: a library for support vector machines,” ACM transactions on intelligent systems and technology (TIST), vol. 2, no. 3, pp. 1–27, 2011.

[24] M. D. Steenwijk et al., “Accurate white matter lesion segmentation by k nearest neighbor classification with tissue type priors (kNN-TTPs),” NeuroImage: Clinical, vol. 3, pp. 462–469, 2013.

[25] H. K. Genant et al., “Comparison of semiquantitative visual and quantitative morphometric assessment of prevalent and incident vertebral fractures in osteoporosis,” Journal of Bone and Mineral Research, vol. 11, no. 7, pp. 984–996, 1996.

[26] H. Weissleder and R. Weissleder, “Lymphedema: evaluation of qualitative and quantitative lymphoscintigraphy in 238 patients,” Radiology, vol. 167, no. 3, pp. 729–735, 1988.

